# Evaluation of gait initiation parameters using inertial sensors in Huntington’s Disease: Insights into anticipatory postural adjustments and cognitive interference

**DOI:** 10.1101/2020.08.13.20174235

**Authors:** Radhika Desai, Nora E. Fritz, Lisa Muratori, Jeffrey M. Hausdorff, Monica Busse, Lori Quinn

## Abstract

**Background and Purpose:** Understanding the contribution of anticipatory postural adjustments (APA) on walking ability in individuals with Huntington’s disease (HD) may provide insight into motor planning and the functional consequences of HD-specific cortical-basal ganglia pathway dysfunctions. The purpose of this study was to evaluate inertial measurement unit (IMU)-derived measures of APAs and first step parameters, and their contribution to gait speed, in individuals with and without manifest HD during a single-task and cognitive load condition.

**Methods:** 33 individuals with manifest HD and 15 age-matched healthy controls wore three Opal APDM IMUs during a 14-meter walk during a single task and cognitive load condition. APA acceleration amplitudes, APA durations, first step range of motion (ROMs), and first step duration were compared, along with their relationship to gait speed.

**Results:** Individuals with HD had significantly greater APA acceleration amplitudes, smaller first step ROMs and longer first step durations compared to healthy controls. No difference in APA durations were present between groups across conditions. Linear model results and significant correlations between mediolateral APA acceleration amplitudes and APA durations were found.

**Conclusions:** Larger acceleration amplitudes, smaller first step ROMs of greater duration, accompanied by the preservation of APA durations reveal a discrepancy in movement scaling in HD. Additionally, the mediolateral component of the APA is likely a rate-limiting factor that drives a compensatory response in gait initiation. Additional research is needed to explore the neural correlates of HD-related movement scaling.

## Introduction

Huntington’s disease (HD) is an inherited neurodegenerative disease that leads to basal ganglia pathway dysfunction of striatal origin, resulting in progressive motor and cognitive disturbances [1]. Gait-related impairments begin in the early stages and worsen through disease progression [2]. Gait initiation, i.e., the preparation and execution of the first step of a gait sequence, reflects the complex integration of movement planning and execution. Anticipatory postural adjustments (APAs) are the first component of gait initiation and are used to stabilize posture in preparation and allowance for lifting the initial stepping leg. APAs are marked by the period of time when the center of mass accelerates laterally and forward, by shifting the center of pressure backward and towards the stance leg just prior to toe-off. Validated accelerometry measures indicate that adequate amplitudes and durations of APAs facilitate first step execution by minimizing balance disturbances [3,4]. Appropriately timed and scaled gait initiation is necessary to maintain stability as an individual transitions from a static posture to dynamic locomotion [5,6].

Individuals with early and mid-stage HD have inadequately scaled APAs and first steps preceding a gait sequence[6]. Specifically, decreased APA displacements and decreased first step durations are evident in individuals with manifest-HD when compared to healthy controls during cued gait initiation [6]. However, durations of APAs are preserved between HD and healthy participants[6]. These findings are in contrast to individuals with Parkinson’s disease (PD) who exhibit both a decrease in APA displacements and decreased APA durations, while they similarly demonstrate a decreased first step length [7]. APA acceleration amplitudes and smoothness, which have also been characterized in PD, provide insight into functional consequences of disease-specific basal ganglia pathway dysfunction, which have not been explored in HD.

Cognitive interference is the relative degradation in motor performance caused by the addition of a cognitive load during a motor task[8,9]. Evaluation of cognitive interference can provide key insights into how attentional and neural resources are distributed. Previous studies have shown that deficits in gait initiation are exacerbated with the addition of an executive functioning task in individuals with PD [10,11]. Similar and yet distinct cortical striatal changes are present in HD, however, the cognitive-motor consequences of these changes have yet to be explored within the context of gait initiation in HD [12].

APAs also play an integral role in overall gait quality, specifically in gait speed and in the maintenance of equilibrium of dynamic stability during step execution. Previous studies [13] have found that mediolateral APAs play a crucial role in controlling mediolateral stability during gait initiation in healthy controls, but this relationship has not been explored in HD. Sufficient motor planning is necessary for the adequate integration of temporal and force parameters of both the anticipatory and voluntary movement, and a mismatch of these parameters may be a reflection of a motor planning deficit.

In this study, we aimed to: 1) determine inertial measurement unit (IMU)-derived measures of APA acceleration amplitudes and durations, and first step range of motion (ROM and durations as previously identified via force plate measures, in individuals with and without manifest HD; 2) evaluate gait initiation parameters under an additional cognitive load in individuals with manifest HD compared to healthy controls; and 3) explore the relationship between APA acceleration amplitudes and smoothness, and the contribution of APA amplitudes to gait speed. We hypothesized IMU-derived measures of APA acceleration amplitudes, but not durations, would be greater and initial step length would be smaller with shorter durations in individuals with manifest- HD when compared to healthy controls. We further hypothesized there would be a larger magnitude of motor deficits of gait initiation parameters when under a cognitive load, as evidenced by increased APA amplitudes and durations, and decreased first step lengths compared to walking while performing an executive function task.

## Methods

### Subjects

Forty-eight participants were recruited for this study as part of the iWEAR-HD project, including 33 individuals with manifest HD (20 males, mean age 54.67 ± 12.57), and 15 healthy controls (8 males, mean age 53.2 ± 13.18). The aforementioned sample size is large enough to indicate gait-related differences in Huntington’s Disease and other rare diseases, as demonstrated by previous gait studies[2,6]. Individuals with HD were recruited from The New York Psychiatric Institute in New York, NY, Wayne State University in Detroit, MI, and the George Huntington Institute in Munster, Germany. Inclusion criteria were: 1) age 18 years or older, 2) diagnosis of HD as confirmed by the individual’s neurologist, 3) capacity to consent, 4) Total Functional Capacity [14] score of 7 or greater, 5) ability to walk 10 meters independently. Exclusion criteria included: 1) diagnosis of juvenile-onset HD, 2) history of co-morbid neurological conditions, 3) acute orthopedic conditions or injuries, 4) inability or unwillingness to give written informed consent (Table 1). The protocol for this study was approved by the Institutional Review Boards of the respective institutions (Teachers College, Columbia University; Wayne State University and the Medical Council Westfalen-Lippe and Westfälische Wilhelms-Universität Münster, on behalf of the George Huntington Institute). All participants signed written informed consent prior to participation.

**Table 1:**
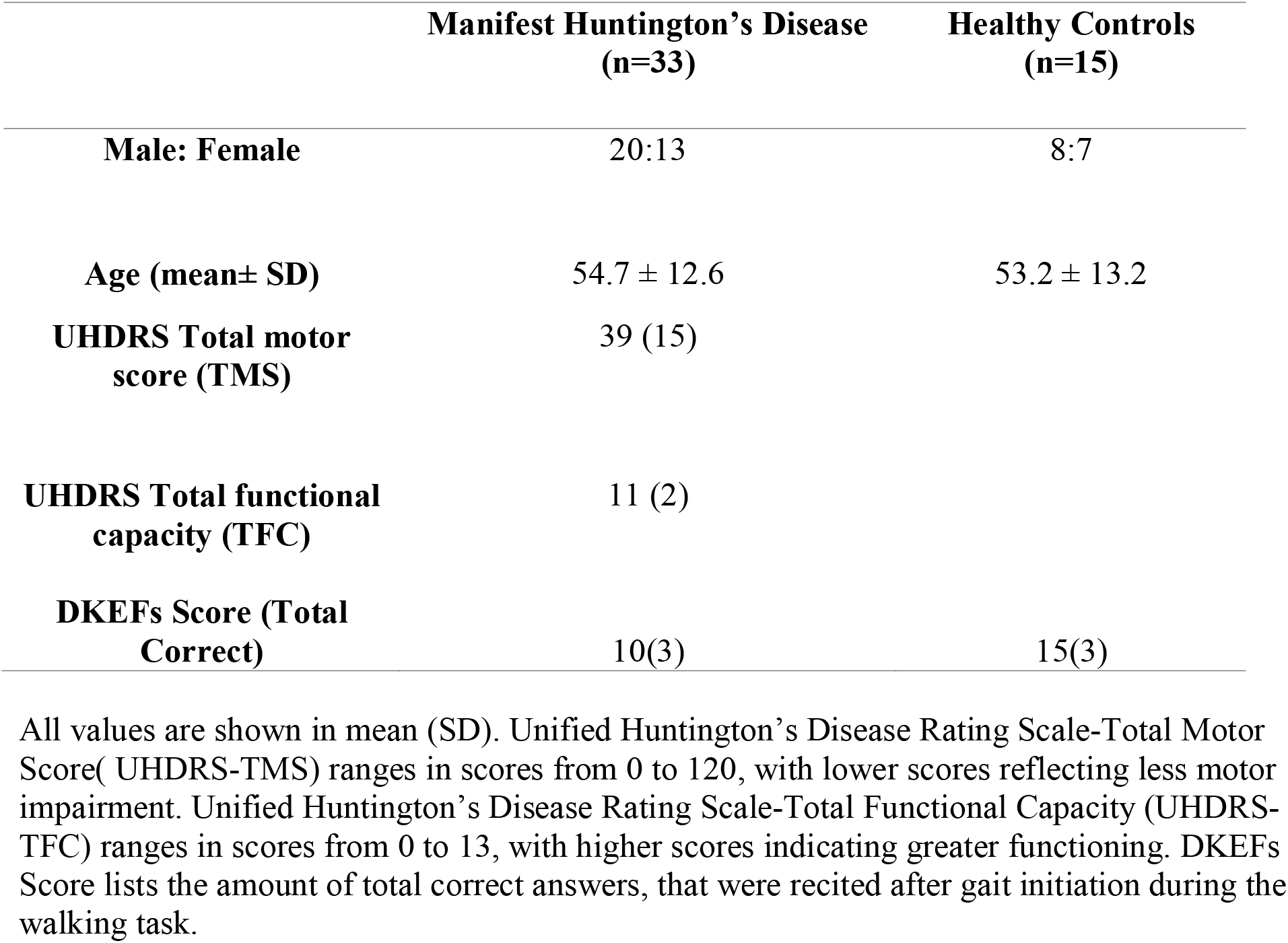
Participant demographics and disease-specific measures in all subjects

### Experimental Setup and Protocol

Opal inertial measurement unit (IMU) sensors (APDM, Inc., Portland, OR) were secured to the dorsum of the feet bilaterally at the level of the metatarsals and the lumbar region of each participant via velcro straps. Participants began a 14-meter walk by standing at a pre-marked location on a level 7-meter walkway. Participants started walking at the sound of a tone, which denoted the synchronization of the APDM sensors and walked to the 7-meter mark, turned and walked back to the start. Synchronization ended once subjects reached the original starting location. The walking condition consisted solely of the 14-meter walk. For the walking condition with an added cognitive load, participants were instructed to begin walking while performing the Delis-Kaplan Executive Function System (D-KEFS) verbal fluency task with category switching. Specifically, participants were asked to alternatively name items of fruit and furniture while walking the same 14-meter distance. Responses were recorded and scored for correct numbers of switches in the walking period as compared to the performance of the D-KEFS category switching test in sitting. Each condition consisted of a single trial with the simple walking task always preceding the more complex walking with the D-KEFS, including the single-task condition of the cognitive task. The D-KEFS has demonstrated reliability and validity in persons with neurologic disorders[15]. Furthermore, the D-KEFS is considered a more complex task of executive functioning and has previously exhibited effects on motor performance [16]. Demographic data including age and sex were collected for all participants. Unified Huntington Disease Rating Scale (UHDRS) measures including Total Functional Capacity (TFC), Total Motor Score (TMS) within 3 months of testing were obtained from the HD participants’ clinical files.

### Extraction of IMU-based measures

Raw signals from the accelerometers were set to horizontal and vertical coordinates by the sensors’ pre-existing algorithms, which utilize the magnetometers and gyroscopes to identify x, y, and z reference positions. The lumbar and ankle accelerometry streams were then extracted and processed in Matlab (Mathworks R2018A). Accelerometry streams, sampled at 128Hz, were filtered through a 3.5Hz cut-off, zero-phase, low-pass Butterworth filter. This filtering profile is consistent with previous uses of APDM IMU sensors in identifying APAs.

APDM Mobility Lab software was unable to apply the standard algorithm for APA processing in most HD participants, secondary to the excessive movements during static stance prior to gait initiation. We, therefore, extracted APAs in a custom Matlab program by first constructing a time window 1000ms prior to time of toe-off of the first stepping leg, as defined as the first peak acceleration in the anterior direction of the ankle sensor. Within this time frame, the maximum value in the mediolateral lumbar accelerometry stream was identified. This value indicated the amplitude of the medial-lateral (ML) APA. The onset of the APA was defined as the time point at which the mediolateral accelerometry stream reaches 20% of the previously identified maximum value within the predetermined time window[17] (Fig. 1). Therefore, the duration of the APA was defined as the length of time between the time of onset to the time of toe-off of the first stepping leg. The anterior-posterior (AP) component of the APA occurs after the first peak of the ML component; amplitudes of the AP component were thus determined by finding the maximum AP value of the lumbar stream within the time window after the max ML peak value and toe-off of the first stepping leg. All accelerometry streams were visually inspected as a second measure to ensure that peak values were determined within appropriate time windows. Duration windows below 10ms and above 7500ms, along with amplitudes below .01(G-units) and above .9(G-units), were not considered as potential APAs[17].

**Fig. 1.**
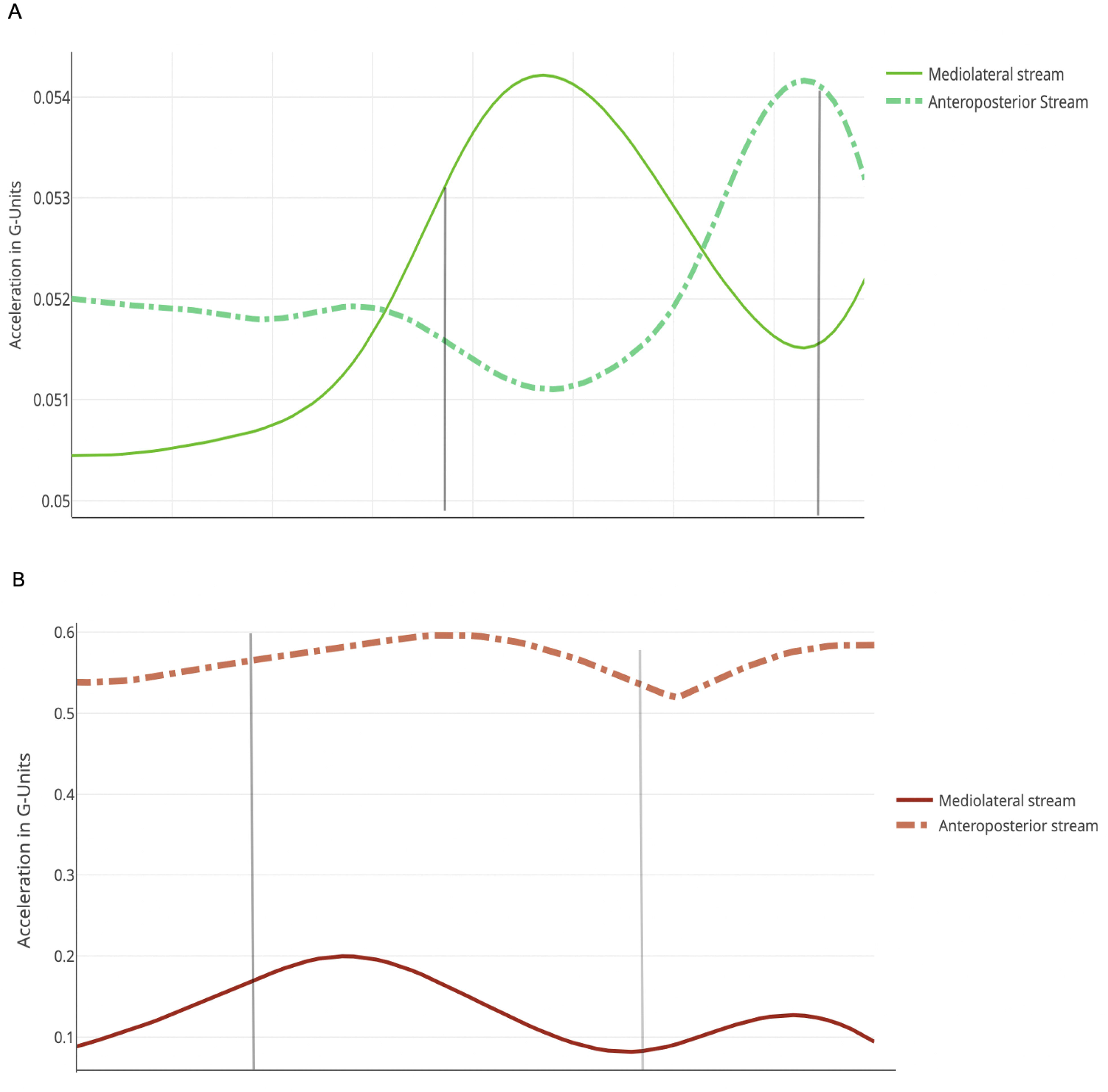
Example acceleration stream from the lumbar sensor of a healthy control and HD participant. Duration of the APA is marked by the grey vertical lines within each accelerometry stream. APA start times are derived from the lumbar stream within a duration window prior to an indicated by toe-off derived from the foot sensor. APA end is the end time indicated by toe-off.

Spatiotemporal parameters of the first step were derived using angular velocities extracted from the ankle gyroscope of the first stepping leg. First step duration was defined as the length of time from APA end to heel strike of the first stepping leg[17]. The time point of APA end was also defined as toe-off. This was identified from the accelerometry data as described above. Specifically, first step duration was defined as the time-to peak angular velocity from APA end to heel strike. The spatial feature of the first step was assessed using the first step range of motion (ROM). ROM was found by integrating the angular velocity of the ankle sensor from APA offset to heel-strike within the gyroscope stream.

Smoothness of the APA in the ML direction was defined as average jerk (m/s^^^3) within the APA period. Specifically, peak acceleration in the ML direction was divided by the duration of the APA. Smoothness of the APA in the AP direction was found in the same manner, using accelerations of the APA in the AP direction as the numerator.

### Statistical Analysis

Statistical analyses were performed in SPSS (SPSS Inc. Released 2018. SPSS for Mac, Version 25.0. Chicago, SPSS Inc). Levene’s test was utilized to assess the homogeneity of variance for amplitudes and durations (p>.05) Gait initiation parameters for the walking alone condition and the walking while performing the D-KEFS condition were analyzed for HD and control groups using a two-way repeated measures ANOVA, with a post-hoc Tukey HSD. Pairwise comparisons of means were used to assess amplitudes and duration across conditions within and between participant groups. A regression analysis was used to determine the degree to which APA parameters could predict gait speed during the 7 meter walk in HD participants. Pearson correlations were utilized to assess correlations between APA amplitudes and APA durations. Signficance value was set at p<0.05 for all analyses.

## Results

Mean values and standard deviations of all outcome measures of groups across task conditions are listed in Table 2. There was a significant main effect (F(1, 46) = 46.65, *p* < .001) for all APA acceleration amplitudes between groups, with HD having greater ML and AP APA acceleration amplitudes(Fig. 2). There were no significant differences in APA durations between groups (p>.05)(Fig. 3). Furthermore, there were no differences found between no-load and cognitive load conditions across all APA parameters in both groups (p>.05).

**Table 2:**
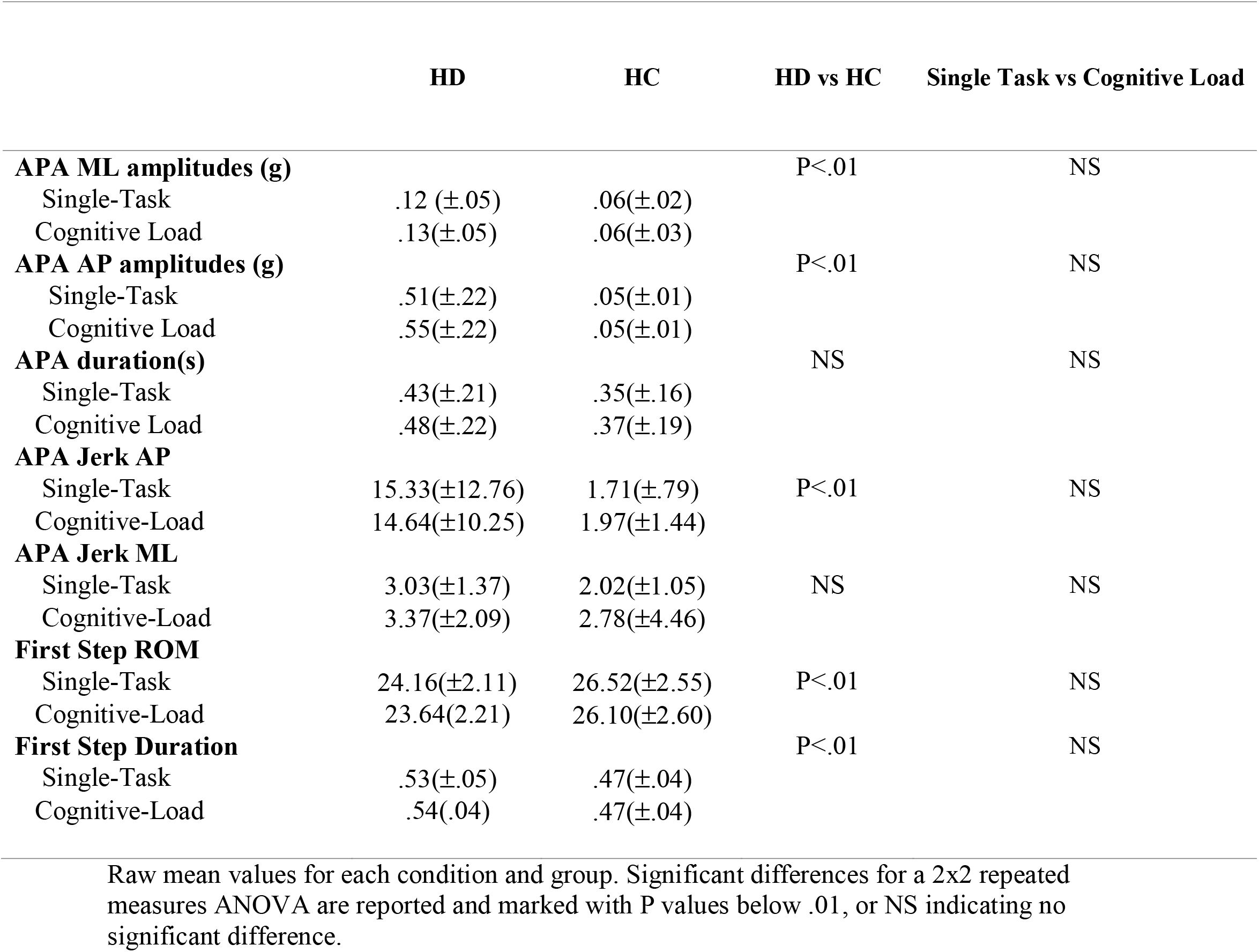
APA values across groups and conditions.

**Fig. 2.**
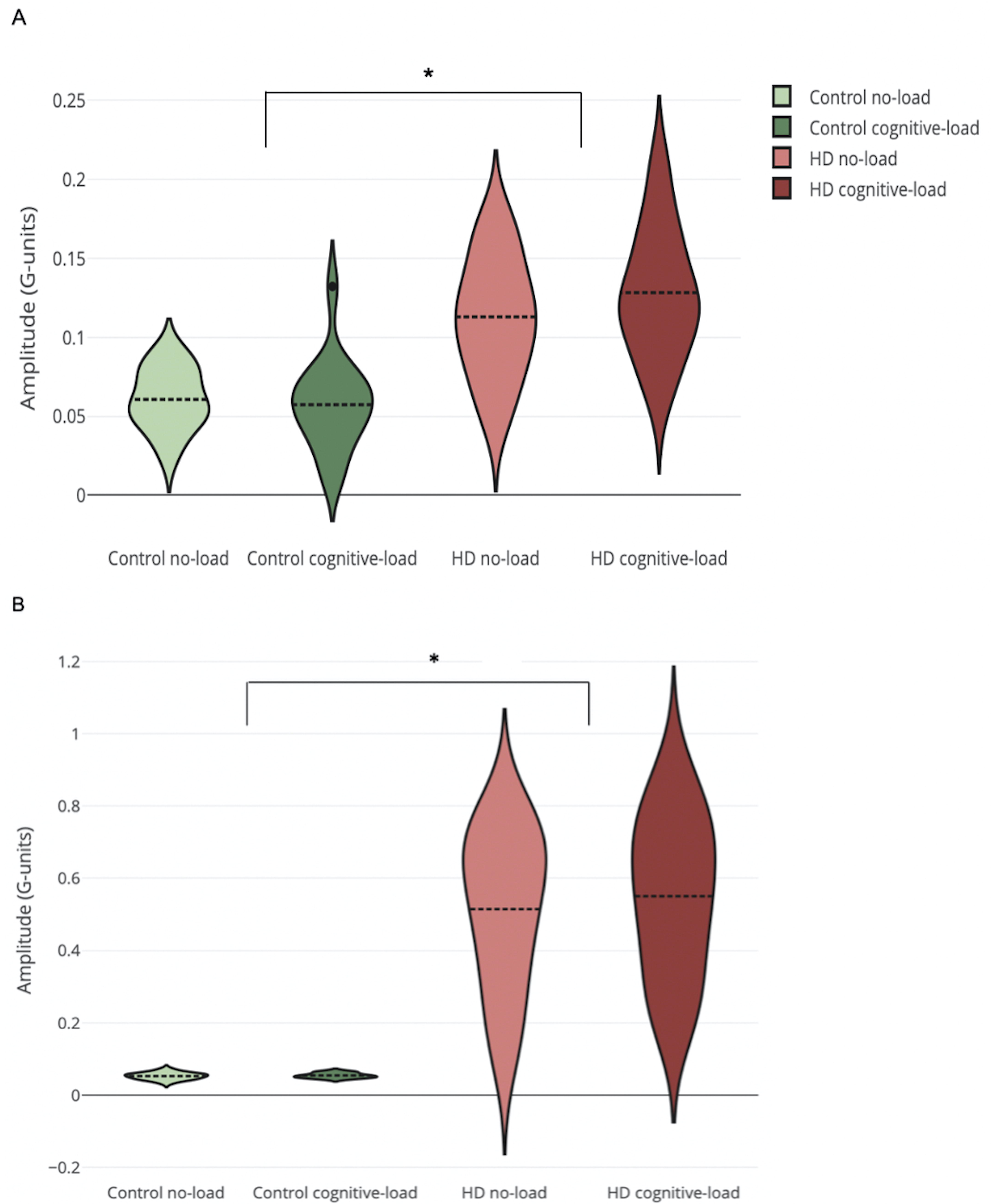
Violin plots display kernel density estimations of APA acceleration amplitudes in the mediolateral (A) and anteroposterior (B) directions. Signifcantly larger accelerations are present between groups for both directions, with drastically larger amplitudes and variance among anteroposterior APAs.

**Fig. 3.**
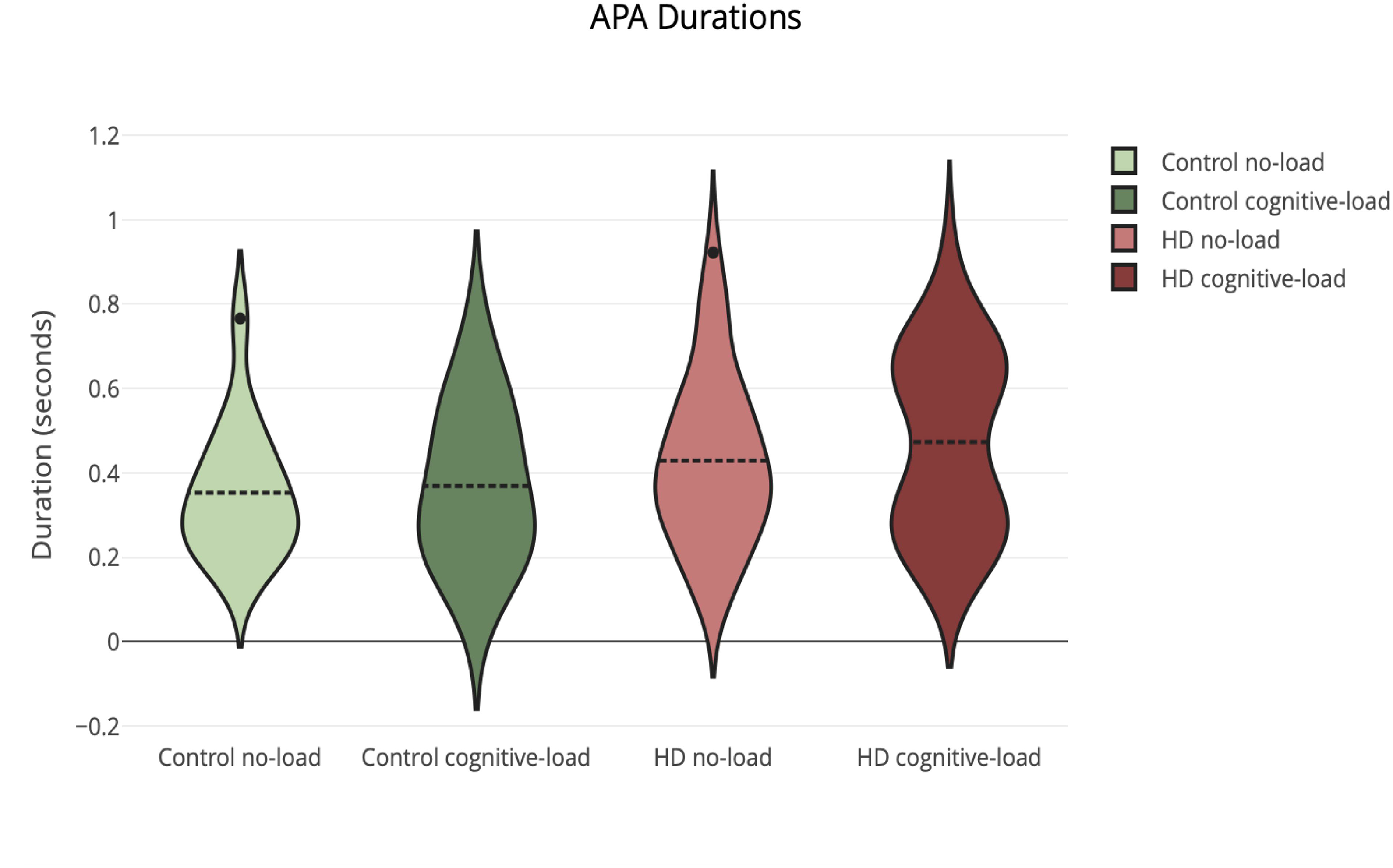
APA durations are preserved across conditions and groups. In contrast to APA amplitude values, variability of durations within each group appear consistent.

The HD group had significantly smaller first step ROM values (F(1,46)=12.67, p=.002) as compared to healthy controls, and larger first step duration values (F(1,46)= 26.20, p=.00) as compared to healthy controls (Fig. 4). There were no differences found between cognitive conditions for first step parameters (p>.05).

**Fig. 4.**
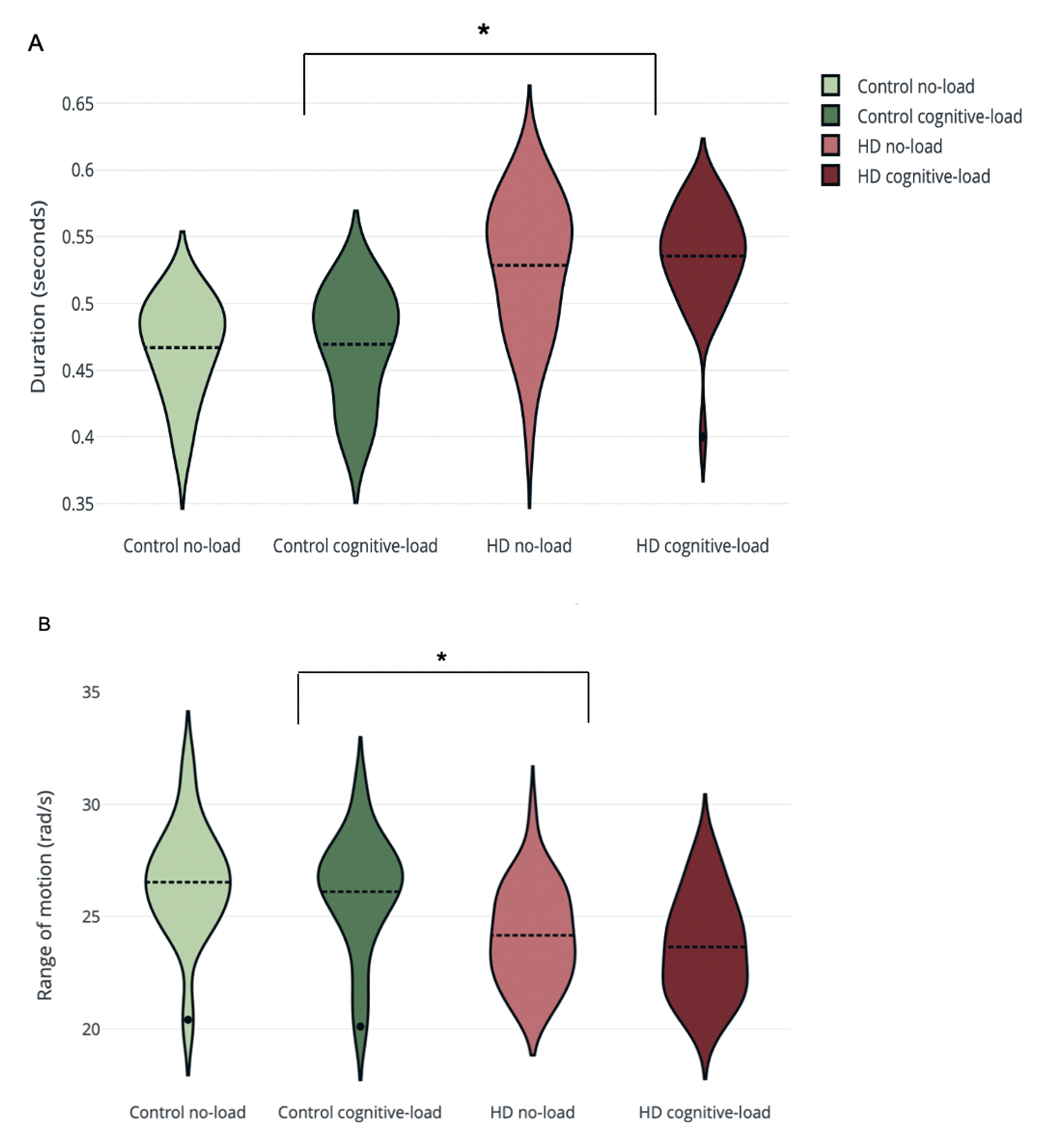
First step durations (A) are larger in the HD group as compared to healthy controls, in contrast to APA durations. The HD group exhibits significantly smaller first step ROM (B) in comparison to healthy controls, however first step range of motion is preserved across conditions.

Among the measures of average jerk during gait initiation, there was a significant difference (F(1,46)=26.05, *p=* .00) of average jerk in the AP direction in the HD group, compared to the healthy control group. However, there was no significant difference in average jerk in the ML direction across groups or conditions (p>.05).

Results of the linear model assessing the role of APA and first step parameters in HD did not indicate a significant model (R^2^ =. 1019, F(4, 26)=1.851, p=.15), however, the mediolateral APA acceleration amplitudes (β = -2.150, p=.04) was a significant predictor of gait speed. Results of a similar analysis among healthy controls found that no APA and first step parameter was a significant predictor of gait speed (p>.05). Lastly, Pearson’s correlations revealed a significant correlation between ML APAs and APA durations among HD individuals(r=.54, p= .002). AP APA amplitudes were not significantly correlated with APA durations. Among the healthy group, neither ML or AP APA amplitudes were significantly correlated with AP duration (p<05).

Results of the repeated measures ANOVA on gait speed during the 7 meter walk indicated a significant main effect for both group (F(1,46)=7.551, p=.01) and condition (F(1,46)= 60.54, p=.00).

## Discussion

Our results confirm previous findings that individuals with HD exhibit greater APA acceleration amplitudes, but not durations, in both the ML and AP directions when compared with healthy controls. Individuals with HD displayed smaller ROM values but remained consistent under a cognitive load. Similarly, the duration of the first steps remained unaffected as a result of cognitive load but was larger in value when compared to healthy controls. The preservation of APA duration between groups may come as a result of inadequate force modulation in the mediolateral direction causing the prioritization of first step completion in the HD group. The HD group had decreased smoothness (greater jerk values) within the AP direction compared to healthy controls. The large correlations between ML APA and APA duration may indicate a dominant rate-limiting factor within gait initiation. The larger accelerations values in HD, with the absence of time scale adjustments, reflects a disintegration between the APA and the associated voluntary movement initiation.

Maintenance of APA duration between groups and conditions with a cognitive load further indicates a mismatch in APA and step initiation since the endpoint of this time window was marked by heel-off (i.e. start of step initiation). This dissociative time scale remained unaltered as a result of cognitive load, indicating preservation despite cognitive interference. This is in contrast to a PD population where first step durations but not APAs are vulnerable to a cognitive load[10]. We speculate that the cognitive task did not interfere with the gait initiation due to motor prioritization and perhaps due to a lack of overlap in the resources required to initiate gait and facilitate working memory in preparation for the recitation of the cognitive task. The lack of motor deficits during cognitive load conditions emphasizes the notion that gait initiation requires a subset of neural pathways and resources distinct from that of continuous gait in HD. This is further emphasized by the results of the dual-task condition on gait speed, which resulted in a significant decrease in speed across groups and conditions.

The mediolateral APA maintains equilibrium as weight is shifted onto the stance leg and is followed by an anteroposterior APA which maintains equilibrium during the first step. As weight is shifted onto the stance leg, deceleration of the body is needed to prevent continuous acceleration, which would otherwise cause destabilizing displacement of the body in the direction of the stance leg. Continuous acceleration towards the stance leg without the appropriate deceleration or a compensatory response could ultimately lead to a lateral fall[13]. In healthy populations, adequate force modulation decelerates the body as it shifts weight onto the stance leg, followed by an acceleration back towards the midline, allowing for smooth backward and forwards anteroposterior APA accelerations through the completion of the first step [13]. However, large and highly variable anteroposterior accelerations among the HD group may reflect compensatory strategy in response to the initial and exceeding mediolateral APA accelerations. A large anteroposterior APA acceleration is an effective compensatory strategy to overcome a lack of mediolateral deceleration and continuous displacement. By accelerating quickly in the AP direction, first step completion is expedited, ultimately preventing dynamic instability and reducing the time spent solely on the stance leg[13]. Furthermore, high correlations between mediolateral APA accelerations and APA durations indicate that the mediolateral APAs dominate the majority of the APA time window and is an essence a rate-limiting factor for gait initiation. Mediolateral APA as a significant predictor of gait speed may also imply that ML shifting is rate-limiting through the entirety of the gait cycle, although we interpret these results of the linear model with caution.

The highly correlated relationship between mediolateral APAs and APA durations, along with exceedingly large and variable AP APAs, argues for a compensatory phenomenon in response to heightened dynamic instability. However, the generalized disintegration of accelerations and durations of the APAs and first step may also come as a result of an HD specific impairment of force modulation, rooted in the disease’s pathophysiology. Generation of the APA and the respective voluntary movement are parallel operations that originate from the basal-ganglia supplementary motor (BG-SMA) complex, and dorsolateral premotor and motor complex respectively[18]. These circuits then meet and integrate within the postural and locomotor centers of the brain stem. However, individuals with HD exhibit hyper-thalamocortical drive within the BG-SMA circuits[19-21].

The consequence of this neurophysiological alteration is an excessive excitatory input into the SMA, putatively causing impairments of force modulation and excessive amplitudes of voluntary movements [20]. Within HD, we speculate that this hyperexcitation of the SMA contributes to deficits in force modulation, and therefore large peak accelerations of APAs. We further posit that hyper-excitation of the SMA contributes to excessive changes in accelerations, which is representative of jerk. Specifically, we quantified this as a value of mean jerk in the ML and AP direction as changes in acceleration over time. HD exhibited larger jerk values in both the ML and AP direction when compared to healthy controls (Figure 4). Our results show that individuals with HD exhibit overall increases in acceleration during a preparatory phase of movement initiation, indicating an impairment of dynamic balance. These large peak accelerations can be explained by the emergence of chorea within a voluntary movement or a standalone deficit in motor planning. As mentioned above, the high correlations between ML APAs and APA durations argue against embedded chorea, since we would not expect to see involuntary movements appropriately scaled and correlated with a time scale of volitional movement. Therefore, it is likely that the predictability of mediolateral APA amplitude on gait speed in individuals with HD is reflective of a disease-specific motor planning deficit.

Spatial-temporal parameters of APAs and the first step are both under cortical and subcortical control. [22]. The supplementary motor area (SMA) is a cortical area highly involved in motor planning, temporal and spatial scaling, and initial motor propagation of the APA. However, the supplementary motor area is highly susceptible to neurophysiological changes as a result of HD progression by means of the basal-ganglia thalamocortical circuit [23]. Due to this susceptibility, increased demands placed on cortical areas that share pathways to the supplementary motor area have the capacity to interfere with motor planning processes, which may disrupt gait initiation in persons with HD.

Whilst it is possible that the cognitive load utilized here may have been insufficiently demanding, future work is needed to further investigate the neural correlates associated with time-scale discrepancies in APAs during gait initiation in individuals with HD. A different task may force reliance on resources and circuitry shared with the SMA, thereby dampening any prefrontal compensatory mechanisms that would normally attenuate hyper-excitation of the premotor areas. Additionally, further research may help to better understand the contextual role of chorea during movements and whether they are enveloped within a voluntary movement.

## Data Availability

Data is available upon request from the authors.

## Acknowledgements

The authors acknowledge the following individuals for assistance with recruitment and data collection at each of the sites: Columbia University: Karen Marder, MD, Paula Wasserman, Deborah Thorne, Ashwini Rao, EdD, OTR, Elizabeth McAneny, and Gregory Youdan. George Huntington Institute: Ralf Reilmann, MD, Stefan Bohlen, MD, Anja Kletsch, Robin Schubert; Wayne State University: Amanda Noakes, Manon Nitta, Mareena Atalla.

The authors also gratefully acknowledge the participants in this study who generously donated their time.

## Declarations

### Ethics approval

The protocol for this study was approved by the Institutional Review Boards of the respective institutions (Teachers College, Columbia University; Wayne State University and the Medical Council Westfalen-Lippe and Westfälische Wilhelms-Universität Münster, on behalf of the George Huntington Institute).

### Consent to participate

All participants signed written informed consent prior to participation.

### Consent for publication

Not applicable

Availability of data and material:

Upon request from the authors

### Authors’ contributions

R.D. contributed to the design of the work, analysis, interpretation of data, and drafted the work. N.E., L.M., and L.Q. contributed to data acquisition, revised the work critically for intellectual content, and approved the version to be published. J.H., M.B. revised the work crtically for intellectual content, and approved the version to be published.

